# Occupational safety and health aspects of corporate social responsibility reporting in Japan: comparison between 2012 and 2020

**DOI:** 10.1101/2022.02.15.22271007

**Authors:** Takahiro Shimizu, Tomohisa Nagata, Ayumi Fujimoto, Shunsuke Inoue, Masako Nagata, Koji Mori

**Author notes:** **Correspondence:** Tomohisa Nagata, MD, PhD, Department of Occupational Health Practice and Management, Institute of Industrial, Ecological Sciences, University of Occupational and Environmental Health, Japan, 1-1, Iseigaoka, Yahatanishi-ku, Kitakyushu 807-8555, Japan.

## Abstract

**Background:** We aimed to survey the content of occupational safety and health (OSH) disclosed in corporate social responsibility (CSR)-related reports and integrated reports in 2020 and to compare the changes between 2012 and 2020 according to industry and company size.

**Methods:** We surveyed the websites of all 2,172 companies listed in the first section of the Tokyo Stock Exchange in 2020 and examined their CSR-related reports and integrated reports. CSR-related reports contain only non-financial information or are posted on webpages addressing sustainability and CSR. Integrated reports include financial information in addition to non-financial information or are posted in the investor relations section. We calculated the proportion of each report type and the proportion of OSH content in each report by industry sector and company size. To increase the reliability of the survey, five experts independently surveyed the same 40 companies and discussed any differing results. We prepared a survey manual based on these discussions. Twenty-five researchers assessed the presence or absence of each type of report and of OSH.

**Results:** Among all companies, 441 (20.3%) companies issued CSR-related reports and 590 (28.1%) issued integrated reports. The number (percentage) of companies that issued either CSR-related reports or integrated reports was 880 (40.5%). The percentages of both CSR-related reports and integrated reports increased with increased company size. The number (percentage) of companies reporting OSH in CSR-related reports was 391 (88.7%) and that in integrated reports was 493 (83.6%). The percentage of OSH reporting in CSR-related reports and integrated reports was high in secondary industries and low in tertiary industries.

**Conclusions:** The companies that issued either a CSR-related report or an integrated report increased by 1.9% in 2020 compared with 2012. The percentage of companies that had described OSH in CSR-related reports and integrated reports were increased in 2020 compared with 2012. Many companies in Japan are emphasizing OSH in their CSR-related and integrated reports.

## Introduction

Corporate social responsibility (CSR) has recently garnered attention and social interest. CSR is defined as a company’s voluntary commitment to social and environmental considerations in the course of its business activities and relationships with stakeholders (all interested parties in organizational activities) [1]. The International Organization for Standardization issued ISO26000 to standardize CSR in 2010 [2]. CSR has become progressively important in social activities that are required to consider sustainability.

The Global Reporting Initiative (GRI) has established sustainability reporting guidelines on how to disclose information on CSR activities, which states that companies are accountable to their stakeholders [3]. CSR-related reports have been used as a medium for information disclosure, and information is disclosed both internally and externally via company websites in the form of PDF files. A previous study pointed out that the number of companies in Japan issuing CSR-related reports had increased as of 2012 [4].

In addition to CSR-related reports, integrated reports have appeared as a method to disclose information to stakeholders, focusing on both non-financial and financial information. This arose owing to the concept of environmental social governance (ESG) proposed by the United Nations through the Principles for Responsible Investment in 2006 and the severe financial crisis triggered by the Lehman Brothers collapse in 2008 [5.6]. This financial crisis increased the need for long-term-oriented corporate management, and the importance of corporate governance and ESG became recognized [5.7]. This prompted the establishment of the International Integrated Reporting Council in 2011, and the International Integrated Reporting Framework was established in 2013 [8]. This framework defined integrated reports as “a concise communication about how an organization’s strategy, governance, performance and prospects, in the context of its external environment, lead to the creation, preservation, or erosion of value over the short, medium, and long term” [8]. The International Integrated Reporting Council has demonstrated that integrated reporting contributes to the stability and sustainability of financial capital markets; this sustainability is a concept that includes not only survival of the companies themselves but also social and environmental issues. The number of companies issuing integrated reports in Japan is reported to be on the increase [8]. It can be inferred that the shift from CSR-related reports to integrated reports is progressing [9, 10].

Occupational Safety and Health (OSH) is included in CSR activities. ISO26000 proposes seven core subjects, one of which is labor practices. Labor practices include OSH, along with respect for the rights of workers, the importance of social dialogue, and responsibility for the human development of workers [2]. The GRI also mentions OSH in GRI403, which lists the specific OSH activities to be disclosed. A survey of Japanese companies listed in the first section of the Tokyo Stock Exchange between 2004 and 2012 reported that an increasing number include OSH in their CSR-related reports [4]. OSH has been emphasized by workers, companies, and society as a factor that contributes to workers, as stakeholders. However, since the emergence of new forms of disclosing information, such as integrated reports, to the best of our knowledge, the methods and descriptions of information disclosure regarding CSR activities and OSH have not been clarified. We aimed to clarify trends in the disclosure of non-financial information and the recognition of OSH since the emergence of ESG and integrated reports by examining changes in the proportions of reports issued and of OSH content described.

We conducted a survey of listed companies to determine the proportions of CSR-related reports and integrated reports published in 2020, and the proportion of content in these reports addressing OSH. We examined changes in the methods of disclosing information on CSR and ESG activities and OSH by company size, classified based on industry and number of employees, comparing the presence of reports and changes in the description of OSH in 2012 and 2020. The purpose of this study was to clarify the trend and development of information disclosure on OSH activities in CSR-related reports and integrated reports.

## Methods

We surveyed all 2,172 companies listed in the first section of the Tokyo Stock Exchange in 2020 to determine the percentage of reports issued and the content of the reports published on each company’s website. The included companies were based on those listed on the website of the First Section of the Tokyo Stock Exchange on August 31, 2020. We surveyed CSR-related reports and integrated reports independently. In cases where both types of report were issued, each report was tabulated separately. The contents of the survey addressed whether a report was issued and whether OSH was described. The distinction between CSR-related reports and integrated reports was determined using the name of the report and location of the report on the website. CSR-related reports can be found in the CSR activities, ESG activities, and sustainability sections of the website. In contrast, integrated reports are often posted in the investor relations section.

If the content of a report comprised only non-financial information, we determined it to be a CSR-related report. However, if financial information was presented together with non-financial information, we judged it to be an integrated report.

The inclusion criteria of a report were that it was published on a website in PDF format or in book form, that the report consisted of six or more pages excluding the front and back covers, and that the report was written in Japanese. The exclusion criteria for the reports were those prepared in English or languages other than Japanese, information disclosed only on the website, and no link to the report.

We assessed the presence or absence of descriptions of OSH activities in each CSR-related report and integrated report. When there was at least one description related to safety or health in the workplace, we judged that there was a description of OSH activities; examples include items related to safety and health, such as efforts to prevent occupational accidents, mental health care, existence of health education, setting of health and safety goals, utilization of occupational safety and health management systems, and the establishment of safety and health committees.

An independent survey of 40 companies was conducted by four experts in occupational medicine and one expert in accounting, making up a total of five researchers [4]. The survey methods were based on those used in previous studies [4]. In the case of differences in the survey results, the causes of the differences were clearly defined and the survey methods were summarized in a manual. From July 2020 to February 2021, a total of 20 people, 16 experts in occupational medicine and four medical students studying occupational medicine, conducted a website survey on the existence of CSR-related reports, integrated reports, and descriptions of OSH. We were unable to examine the contents of all integrated reports during this period. Therefore, from July to September 2021, five medical students conducting research in occupational medicine carried out an additional survey of the integrated reports. In this case, we surveyed reports published in 2020 and excluded those published in 2021. Each researcher surveyed the same companies. In cases where the results of the survey were not matched (for example, presence or absence of reports, categorized types of report, and OSH mentioned or not), we examined the causes and made modifications to standardize the survey.

## Statistical analysis

We classified the companies into 10 industries, as in a previous study, to determine the percentage of reports issued by industry and company size and the proportion of OSH. We categorized company size according to the number of employees. The 10 industries were Fisheries, agriculture, and forestry; Mining; Construction; Manufacturing; Electricity and gas; Transportation, information, and communication; Commerce; Finance and insurance; Real estate; and Services. The number of employees was classified into ranges of 49 or fewer, 50–299, 300–999, 1000–2999, 3000–4999, 5000– 9999, and 10,000 or more; the percentage of reports issued was investigated.

We examined the relationship among company size according to industry, number of employees, and percentage of reports issued, type of report, and descriptions of OSH. Statistical analysis was performed using Stata/SE 16 software for Windows (Stata Corp LLC, College Station, TX, USA).

We only used publicly available information in the study.

## Results

Table 1 shows the number of listed companies by industry and company size based on the number of employees. In 2020, the number of listed companies was 2172. By industry, the Manufacturing sector was the largest with 914 companies (42.1%), followed by Commerce with 381 companies (17.5%), Transportation, information, and communication with 305 companies (14.0%), and Services with 225 companies (10.4%). By size, companies with 300–999 employees accounted for the largest number of companies with 756 (34.8%), and the number of companies in each category decreased with increased company size.

**Table 1.** Description of Japanese companies listed in the first section of the Tokyo Stock Exchange.

|  | 2012 |  | 2020 |  |
| --- | --- | --- | --- | --- |
|  | N | % | N | % |
| Total | 1717 |  | 2172 |  |
| Industry category |  |  |  |  |
| Fisheries, agriculture, and forestry | 5 | 0.3 | 7 | 0.3 |
| Mining | 7 | 0.4 | 6 | 0.3 |
| Construction | 97 | 5.6 | 102 | 4.7 |
| Manufacturing | 844 | 49.2 | 914 | 42.1 |
| Electricity and gas | 17 | 1.0 | 22 | 1.0 |
| Transportation, information, and communication | 169 | 9.8 | 305 | 14.0 |
| Commerece | 300 | 17.5 | 381 | 17.5 |
| Finance and insurance | 129 | 7.5 | 139 | 6.4 |
| Real estate | 48 | 2.8 | 71 | 3.3 |
| Services | 101 | 5.9 | 225 | 10.4 |
| Company size |  |  |  |  |
| –49 | 70 | 4.1 | 139 | 6.4 |
| 50–299 | 274 | 16.0 | 462 | 21.3 |
| 300–999 | 594 | 34.6 | 756 | 34.8 |
| 1000–2999 | 501 | 29.2 | 511 | 23.5 |
| 3000–4999 | 133 | 7.7 | 126 | 5.8 |
| 5000–9999 | 69 | 4.0 | 80 | 3.7 |
| 10000– | 59 | 3.4 | 57 | 2.6 |
| uncertain | 17 | 1.0 | 41 | 1.9 |

N: Number of companies.

Table 2 describes CSR-related and integrated reports issued in 2020. The number (percentage) of companies that issued CSR-related reports was 441 (20.3%), and integrated reports were issued by 590 companies (28.1%); the proportion of integrated reports was higher than that of CSR-related reports. In total, 880 companies (40.5%) issued CSR-related reports or integrated reports.

**Table2.**
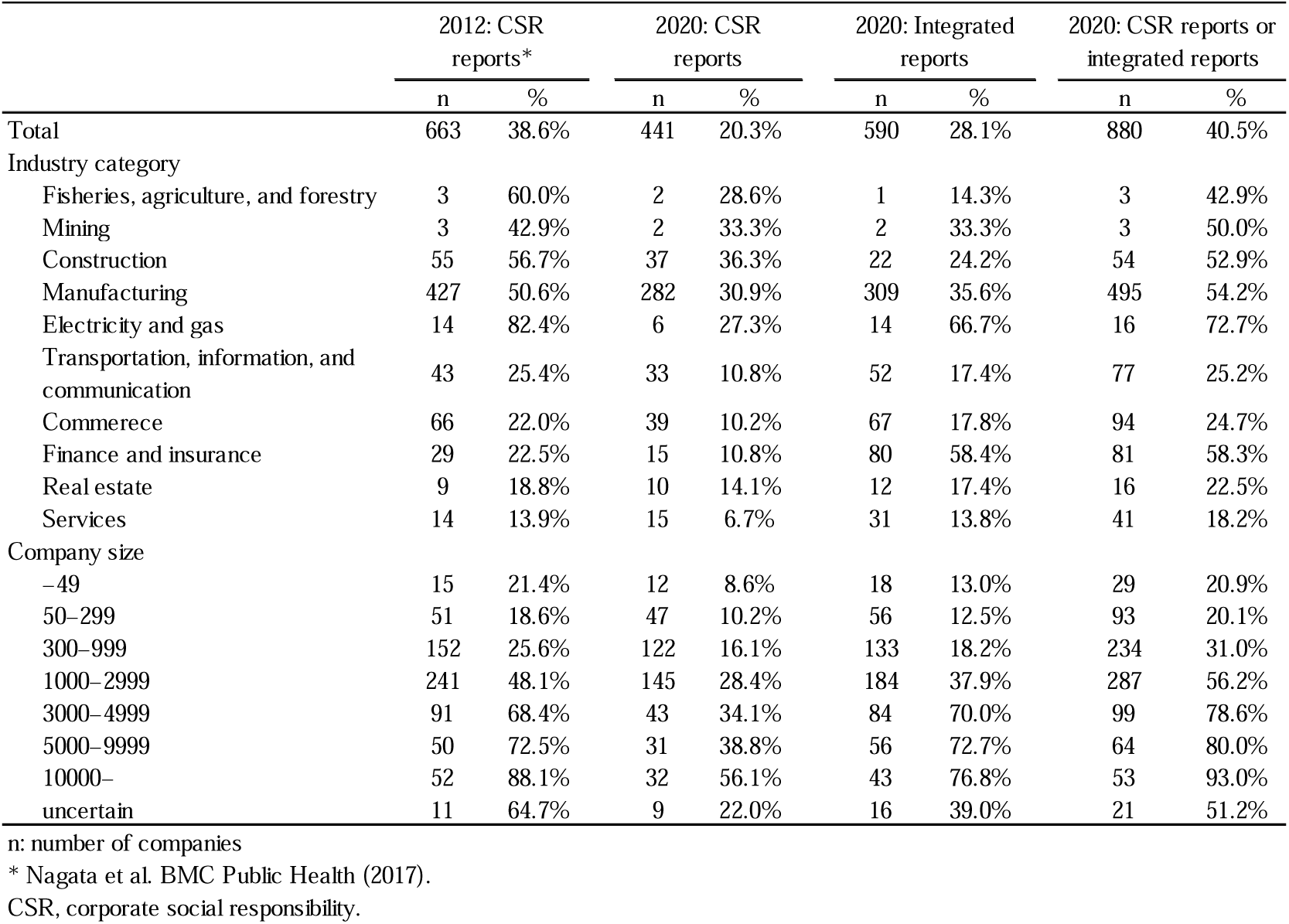
Companies listed in first section of Tokyo Stock Exchange with published CSR-related and integrated reports.

In 2020, by industry, 37 companies (36.3%) in the Construction sector issued the highest percentage of CSR-related reports, followed by 2 companies (33.3%) in the Mining sector and 282 companies (30.9%) in the Manufacturing sector. The number (percentage) of companies issuing integrated reports was highest in the Electricity and gas sector at 14 (66.7%), followed by the Finance and insurance sector at 80 (58.4%) and the Manufacturing sector at 309 (35.6%). Among companies that issued either a CSR-related report or an integrated report, 16 (72.7%) companies in the Electricity and gas category issued the highest percentage of reports, followed by 81 (58.3%) in the Finance and insurance category, 495 (54.2%) in the Manufacturing category, 54 (52.9%) in the Construction category, and 3 (50.0%) in the Mining category.

The percentage of published CSR-related reports increased consistently with increased company size in 2020. For integrated reports, there was a large increase from 18.2% to 37.9% in companies with 300–999 and 1000–2999 employees, respectively, and a further increase to 70.0% in the next tier of 3000–4999 employees. The percentage of companies that issued either CSR-related reports or integrated reports increased with increased company size, and the percentage increased sharply from 1000–2999 to 3000–4999 employees.

Table 3 presents descriptions of OSH activities in CSR-related reports and integrated reports in 2020. The rate of OSH descriptions was 88.7% in CSR-related reports and 83.6% in integrated reports during 2020. By industry, 100% of companies in the Electricity and gas and in the Fisheries, agriculture and forestry sectors, 94.6% in the Construction sector, 91.5% in the Manufacturing sector, and 90.0% in the Real estate sector described their OSH activities in their 2020 CSR-related reports. The percentage of OSH activities described in integrated reports was more than 83.6% in Fisheries, agriculture, forestry; Mining; Construction; Electricity and gas (100%), and Manufacturing (90.3%).

**Table 3.** Companies describing occupational safety and health activities in CSR-related or integrated reports.

|  | 2012: CSR reports* |  | 2020: CSR reports |  | 2020: Integrated reports |  |
| --- | --- | --- | --- | --- | --- | --- |
|  | n(OSH) | % *1 | n(OSH) | % *1 | n(OSH) | % *2 |
| Total | 507 | 76.5% | 391 | 88.7% | 493 | 83.6% |
| Industry category |  |  |  |  |  |  |
| Fisheries, agriculture, and forestry | 2 | 66.7% | 2 | 100.0% | 1 | 100.0% |
| Mining | 3 | 100.0% | 1 | 50.0% | 2 | 100.0% |
| Construction | 47 | 85.5% | 35 | 94.6% | 22 | 100.0% |
| Manufacturing | 337 | 78.9% | 258 | 91.5% | 279 | 90.3% |
| Electricity and gas | 12 | 85.7% | 6 | 100.0% | 14 | 100.0% |
| Transportation, information, and communication | 37 | 86.0% | 27 | 81.8% | 43 | 82.7% |
| Commerce | 42 | 63.6% | 27 | 69.2% | 49 | 73.1% |
| Finance and insurance | 14 | 48.3% | 13 | 86.7% | 53 | 66.3% |
| Real estate | 7 | 77.8% | 9 | 90.0% | 10 | 83.3% |
| Services | 6 | 42.9% | 13 | 86.7% | 20 | 64.5% |
| Company size |  |  |  |  |  |  |
| –49 | 12 | 80.0% | 7 | 58.3% | 12 | 66.7% |
| 50–299 | 40 | 78.4% | 37 | 78.7% | 46 | 82.1% |
| 300–999 | 101 | 66.4% | 109 | 89.3% | 99 | 74.4% |
| 1000–2999 | 182 | 75.5% | 128 | 88.3% | 156 | 84.8% |
| 3000–4999 | 74 | 81.3% | 39 | 90.7% | 76 | 90.5% |
| 5000–9999 | 42 | 84.0% | 31 | 100.0% | 53 | 94.6% |
| 10000– | 47 | 90.4% | 31 | 96.9% | 37 | 86.0% |
| uncertain | 9 | 81.8% | 9 | 100.0% | 14 | 87.5% |

n (OSH): number of companies that describing OSH in reports.
\* Nagata et al. BMC Public Health (2017).
\*1: proportion of companies describing occupational safety and health activities among companies that published CSR-related reports.
\*2: proportion of companies describing occupational safety and health activities among companies that published integrated reports.
CSR, corporate social responsibility; OSH, occupational safety and health.

With regard to company size, the percentage of companies with more than 3000–4999 employees and that published CSR-related reports was more than 88.7% of the total CSR-related reports in 2020. In the case of published integrated reports, the percentage of companies with more than 1000–2999 employees exceeded 83.6% of all integrated reports.

## Discussion

We examined the method of disclosing information on CSR and ESG activities in comparison with a previous study in 2012 and found that the number of CSR-related reports published by companies in Japan has decreased and the percentage of any reports, including integrated reports, has increased slightly. Additionally, we found that the publication of reports and description of OSH are unevenly distributed among industries and companies of different sizes. We discuss these factors and future prospects below.

The percentage of CSR-related reports was decreased compared with this percentage in 2012. This suggests that with the advent of ESG, some companies that formerly issued CSR-related reports have switched to issuing integrated reports. Integrated reports appeared with the emergence of the Integrated Reporting Framework of the IIRC in 2013 [8]. We considered integrated reports to be similar to CSR-related reports, which deal with non-financial information, and we assessed whether any of these two types of report had been issued by companies [8]. The percentage of any of these reports issued increased slightly, indicating that the rising trend of issuing reports is ongoing.

How OSH is disclosed in CSR-related reports and integrated reports is based on the assumption that companies issue each report. We first checked the publication status of each report by industry and company size. Secondary industries represented by Electricity and gas, Manufacturing, and Construction both had a high percentage of CSR-related reports and integrated reports issued in 2020; the same trend was observed in 2012 [4]. Additionally, the proportion of secondary industries that issued integrated reports was high, indicating that these companies are now issuing more integrated reports. The percentage of CSR-related reports issued by companies in the Electricity and gas sector decreased significantly, and that of integrated reports in this sector increased the most. Electricity and gas showed the largest shift in the form of report publication compared with 2012.

Tertiary industries such as Services; Real estate; Commerce; and Transportation, information, and communication had low rates of issuing CSR-related and integrated reports in 2020. In particular, the service industry had the lowest rate of issuing CSR-related reports and integrated reports, both in the current study and in a previous study [4]. We considered that some tertiary industries are not interested in publishing these reports. However, the publication rate of CSR-related reports in 2020 was low in the Finance industry as well as in other tertiary industries, but a high publication rate was observed for integrated reports in this sector. This is because banks are required to issue periodic reports according to Article 21 of the Banking Act in Japan; these periodic reports are called integrated reports or annual reports, in which non-financial information is disclosed [11].

In terms of company size according to number of employees, the percentages of CSR-related reports and integrated reports issued increased with the size of the company. The same trend was observed for CSR-related reports in 2012 [4]. This indicates that the larger the company size, the more active it is in issuing integrated reports. It has been reported that the ESG score provided by rating agencies, used to evaluate the ESG management of a company, is positively correlated with the size of the company [12]. Larger companies are more active in ESG activities and may have a higher percentage of non-financial information that is disclosed and a higher ESG score.

When considering the purpose of each type of report, the percentage of OSH content in CSR-related reports in 2020 increased, suggesting that OSH is becoming increasingly important. The percentage of OSH in integrated reports was also high, suggesting that not only companies but also investors and stakeholders are interested in OSH. The percentage of OSH described in CSR-related reports and integrated reports suggests that OSH may be more strongly recognized as a social responsibility.

The percentage of OSH in CSR-related reports in 2020 was more than 80% in all industries except Commerce and Mining. The percentage of OSH in the tertiary sector was low for integrated reports. This suggests that OSH is recognized as a social responsibility and comprises non-financial information in a wide range of industries, although it is considered that tertiary industries such as Commerce have less recognition that OSH is information that should be disclosed to stakeholders. A program exists to survey and certify the health promotion efforts of employees in Japan, but the participation rate of tertiary industries in this health management survey is low [13]. It is considered that tertiary industries may have low awareness about OSH. OSH includes the element of safety, so it is possible that tertiary industries, which do not directly perform hazardous work, do not have sufficient descriptions about safety, resulting in low results. Additionally, companies are required to have high awareness about human rights throughout the supply chain [14]. Companies within the supply chain may also be required to have a high level of awareness about human rights, and this may be a factor that affects the percentage of OSH content in reports.

The percentage of OSH in CSR-related reports in 2020 increased with increased company size. For integrated reports, the percentage of companies with 1000– 2999 employees or more exceeded the overall percentage of companies that issued integrated reports. The larger the company, the higher the proportion of OSH content in the integrated report. It is thought that this is because the larger the company, the more resources it has to promote OSH and the greater its responsibility to society to promote health and safety. In CSR-related reports and integrated reports for 2020, the proportion of OSH descriptions in the reports was lowest for companies with 49 or fewer employees. According to the law in Japan, the health and safety system changes depending on whether there are 50 or more employees, [15], which may have an effect on the rate of reporting on health and safety. Specifically, the obligation to hold health and safety committee meetings, to appoint a health and safety promoter, and to appoint an industrial physician arises when the number of employees exceeds 50, which may cause differences in health and safety efforts [15]. Although it is not possible to generalize the results of this survey because there is a possibility that the health and safety department exists separately from the holding company, it is considered a factor to be noted.

It is likely that the publication format of reports will continue to change from that in this survey. Whether the shift from CSR-related reports to integrated reports will progress needs to be investigated in future surveys. In this survey, the percentage of OSH entries was examined, but factors influencing the proportion of OSH content was not clarified. However, the frequency was higher in tertiary industries compared with secondary industries, according to Japanese statistics regarding industrial accidents [16]. Companies in the tertiary industry sector and other industries with low OSH reporting may be able to contribute to improve OSH for workers by promoting OSH disclosure.

There are at least three strengths in this paper. First, we conducted a survey of all listed companies in 2020. Second, some studies have investigated the changes in the number of integrated reports issued in Japan; however, to our best knowledge, no studies have investigated the changes in CSR-related reports and integrated reports. Third, to the best of our knowledge, no studies have observed the rate of OSH in CSR-related reports and integrated reports. From the results of this study, it is possible to observe changes in the nature of disclosure of non-financial information in Japan and the extent to which OSH is given attention in such disclosure, which may provide a basis for health and safety professionals to call for the incorporation of OSH in ESG and CSR-related activities.

There are at least three limitations of this survey. First, variation among surveyors cannot be completely excluded. We tried to eliminate the differences in understanding and perception among researchers by developing a manual based on past survey methods, involving all researchers in the conduct of surveys within the same company, and examining and defining differences in the results. However, we cannot say that we were able to completely exclude these differences; there is a possibility that there were differences in judgment among the researchers. Second, there were 75 integrated reports written only in English, which were excluded from the survey. An integrated report is a medium for disclosing non-financial information to investors, including those who are overseas, and it differs from the nature of CSR-related reports; this is thought to have caused differences in the language of disclosure. Therefore, the publication rate of reports and the rate of OSH disclosure may vary from the figures in this survey. However, the number of integrated reports issued by listed companies in 2012 was very small (57 companies); we consider that this did not have a significant impact on the results of the 2012 survey. Additionally, the number of reports in English was small compared with the total number of reports, and the analysis was conducted after excluding the number of integrated reports “issued”, so we believe that the impact is small. Third, if the number of companies that directly list CSR/ESG information on their websites increases, this survey may not accurately represent the actual status of non-financial information disclosure. However, standardizing the condition that the survey be in the form of a report offers the advantage of being able to make comparisons with past surveys. The accounting firm KPMG reports that the number of integrated reports is increasing each year. Because information disclosure in the form of a report is still considered to be the norm, we believe it is appropriate to conduct a survey focusing on reports [9.10].

## Conclusions

There was a slight increase in the percentage of companies that issued either CSR-related reports or integrated reports in 2020, as compared with 2012. The percentage of companies issuing integrated reports has been increasing. In particular, the percentage of disclosed OSH-related content also increased in both CSR-related reports and integrated reports. The medium for disclosure of non-financial information is changing to integrated reports with the emergence of ESG, and recognition of the importance of OSH is continuously improving. It is expected that research and comparisons of integrated reports will become increasingly important in the future.

## Data Availability

All data produced in the present study are available upon reasonable request to the authors.

## Abbreviations

CSR: Corporate social responsibility
ESG: Environmental social governance
OSH: Occupational safety and health
GRI: Global Reporting Initiative

## Declarations

### Ethics approval and consent to participate

All data used for the study are publicly available.

### Consent for publication

Not applicable.

### Availability of data and materials

Data supporting these findings are stored by Occupational Health Practice and Management, Institute of Industrial Ecological Sciences, University of Occupational and Environmental Health, Japan. Requests for information should be directed to the corresponding author.

### Authors’ contributions

TS, TN and KM planned the study. TS and TN drafted the manuscript. TS, TN, AF, SI and MN created the database of CSR reporting and Integrated reporting. TS and TN performed the statistical analysis. All the authors read and approved the final manuscript.

## Acknowledgments

The authors express their sincere appreciation to Dr. Masanao Kaneto of Hosei University and Dr. Takahiro Mori, Dr. Miho Omori, Dr. Kosuke Sakai, Dr. Hirosuke Takahashi, Dr. Kotaro Nagata, Dr. Masatoshi Goami, Dr. Hiroyuki Kuwabara, Dr. Naozumi Sueyoshi, Dr. Hayato Shimoda, Dr. Riku Hachisuka, Dr. Ryoma Kin, Dr. Mio Shibagaki, Dr. Satoru Fujisawa, Dr. Natsuho Miura, Dr. Haruka Takahata, Dr. Ryo Ishimaru, Dr. Souma Shirasawa, Dr. Kakeru Tsutsumi, and Dr. Tomoya Sakai of the University of Occupational and Environmental Health, Japan, who supported us in the analysis of CSR-related reporting and integrated reports. We thank Analisa Avila, MPH, ELS, of Edanz (https://jp.edanz.com/ac) for editing a draft of this manuscript.

## References

1. Zwetsloot G, Starren A. European Agency for Safety and Health at work: corporate social responsibility and safety and health at work. Luxembourg: Office for Official Publications of the European Communities; 2004.

2. International Organisation for Standardization. Guidance on social responsibility (ISO 26000:2010) https://www.iso.org/iso-26000-social-responsibility.html. Accessed 7 Feb 2022.

3. Global Reporting Initiative: G4 sustainability reporting guidelines. 2013. https://respect.international/wp-content/uploads/2017/10/G4-Sustainability-Reporting-Guidelines-Implementation-Manual-GRI-2013.pdf. Accessed 7 Feb 2022.

4. Tomohisa N, Akinori N, Koji M, Takashi M, Futoshi K, Masako N. Occupational safety and health aspects of corporate social responsibility reporting in Japan from 2004 to 2012. BMC Public Health. 2017;17:381.

5. Principles Responsible Investment. About the PRI. https://www.unpri.org/pri/about-the-pri. Accessed 7 Feb 2022.

6. Miralles-Quirós M, Miralles-Quirós J, Hernández, J. ESG performance and shareholder value creation in the banking industry: international differences. Sustainability. 2019;11:1404.

7. International Integrated Reporting Council. 10 years of the IIRC. https://www.integratedreporting.org/10-years/10-years-summary. Accessed 7 Feb 2022.

8. International Integrated Reporting Council. International <IR> Framework. https://www.integratedreporting.org/resource/international-ir-framework/. Accessed 7 Feb 2022.

9. KPMG Japan. Corporate Governance Center of Excellence Integrated Reporting Task Force. Survey of Integrated Reporting in Japan 2020. https://assets.kpmg/content/dam/kpmg/jp/pdf/2021/jp-integrated-reporting2020.pdf. Accessed 7 Feb 2022.

10. Corporate Value Reporting Lab. Trends in Integrated Reporting that Support Sustainable Growth in Japan 2020. http://cvrl-net.com/archive/pdf/list2020_en_202105.pdf. Accessed 7 Feb 2022.

11. Banking Act. Article 21 (1). https://elaws.e-gov.go.jp/documentãlawid=356AC0000000059. Accessed 7 Feb 2022.

12. Samuel D, Christian K, Bernhard Z. The influence of firm size on the ESG score: corporate sustainability ratings under review. J Bus Ethics. 2020; doi:10.1007/s10551-019-04164-1.

13. Ministry of Economy, Trade and Industry, Japan. Survey of health management level. Promotion of Health Management, and regarding “Health Management Brand 2021” and “Excellent Health Management Corporation 2021”. https://www.meti.go.jp/policy/mono_info_service/healthcare/downloadfiles/1_METI_R2kenkoukeieikensyoseido_setsumei_shiryo.pdf. Accessed 7 Feb 2022.

14. United Nations. Guiding Principles on Business and Human Rights.Implementing the United Nations “Protect, Respect and Remedy” Framework. 2022.

15. Ordinance on Industrial Safety and Health, Japan. https://www.ohchr.org/documents/issues/business/a-hrc-17-31_aev.pdf. Accessed 7 Feb http://www.japaneselawtranslation.go.jp/law/detail/ãid=3878&vm=&re=. Accessed Feb 2022.

16. Ministry of Health, Labour and Welfare, Japan. Survey of Occupational Acciden Trends (Survey of Workplaces with 100 or More Employees and General Construction Industry) in Japan. https://www.mhlw.go.jp/toukei/itiran/roudou/saigai/20/dl/2020toukeihyo.pdf. Accessed 7 Feb 2022.

